# An AI Model Identifies Chemotherapy Benefit in Node-Negative HR+/HER2-Breast Cancer Patients from TAILORx, a Phase 3 Randomized Clinical Trial

**DOI:** 10.64898/2026.09.12.26362500

**Authors:** Nancy Chan, Cerise Tang, Dhruva Biswas, Jungkyu Park, Ken G. Zeng, Joseph Cappadona, Chuwen Liu, Alec McClean, Jeroen Berrevoets, Linus Bao, Bartosz Machura, Robert J. Gray, Victoria Wang, Jan Witowski, Krzysztof J. Geras, Joseph A. Sparano, TAILORx Investigators

## Abstract

**BACKGROUND:** The prescribing of adjuvant chemotherapy for patients with breast cancer must balance the survival benefit against treatment-related toxicities. TAILORx, a phase 3 randomized clinical trial, demonstrated that endocrine therapy alone was, on average, noninferior to chemoendocrine therapy for patients classified as intermediate risk by a 21-gene Recurrence Score (RS) assay. Uncertainty remains about the optimal strategy to identify which intermediate-RS individuals derive benefit from chemotherapy.

**METHODS:** We analyzed 6735 patients from the TAILORx trial using Ataraxis Breast CTX with pathology images (H&E slides) and RS available. CTX is an AI model which integrates morphologic features extracted from H&E-stained slides with clinical variables to generate individualized predictions of prognosis (CTX-prognostic) and chemotherapy benefit (CTX-benefit). No data from TAILORx were used to train CTX and CTX was not retrained or recalibrated for this study. Analyses were prespecified in a protocol approved by ECOG-ACRIN.

**RESULTS:** Among patients treated with endocrine therapy alone, CTX-prognostic predicted disease-free interval (DFI), with a hazard ratio per 1 SD increase of 1.651 (95% CI, 1.511-1.803, p < 0.001) and C-index of 0.736 (95% CI, 0.697-0.770). Performance was similar in patients treated with chemoendocrine therapy (hazard ratio = 1.642 [95% CI, 1.495-1.804, p < 0.001], C-index = 0.720 [95% CI, 0.681-0.756]). After adjusting for the RS, CTX-prognostic remained significantly associated with DFI in both treatment groups. In the intermediate-RS subgroup, the treatment-by-biomarker interaction for CTX-benefit was significant (p = 0.016), with patients identified as high-benefit deriving a 5.6% increase in observed 5-year DFI rates from the addition of chemotherapy, compared with no detectable benefit in low-benefit patients. Stratifying the same subgroup by RS at a threshold selecting a similar proportion of patients yielded no significant interaction (p = 0.20).

**CONCLUSIONS:** In an analysis of TAILORx, CTX-prognostic stratified patients according to their risk of recurrence. Among intermediate-RS patients, CTX-benefit identified who derived clinically meaningful benefit from adjuvant chemotherapy, a population currently not identifiable by the RS alone.

## Introduction

Hormone receptor-positive, human epidermal growth factor receptor 2-negative (HR+/HER2-) disease accounts for approximately 70% of cases.^1^ Randomized clinical trials have established that adding chemotherapy to endocrine therapy reduces recurrence,^2,3^ but the magnitude of benefit varies substantially between individuals.^4^ Overtreatment exposes patients to severe acute and long-term toxicities, financial burden, and reductions in quality of life,^5–7^ whereas undertreatment risks preventable recurrence.

Current guidelines advise using genomic risk scores to inform adjuvant chemotherapy decision-making. The 21-gene Recurrence Score (RS) assay was prognostic for distant recurrence.^8^ Moreover, patients with high RS (>26) derived benefit from the addition of chemotherapy, whereas patients with low-RS (<11) had low rates of distant recurrence with endocrine therapy alone and derived little benefit from chemotherapy.^9,10^ Whether patients with intermediate-RS (11-25) derived clinical benefit of chemotherapy remained uncertain, and thus, the TAILORx trial randomly assigned patients with RS 11-25 to either endocrine therapy alone or chemoendocrine therapy. The trial demonstrated noninferiority of endocrine therapy alone in the intermediate-RS population and therefore, was not able to identify patients with intermediate-RS who may benefit from adjuvant chemotherapy.^11^

Existing assays, including the RS, were developed to estimate recurrence risk.^8,12^ Although higher scores have been associated with greater average chemotherapy benefit,^10,11^ RS itself does not estimate an individual patient’s reduction in recurrence risk with the addition of chemotherapy. Rather than deriving a prognostic score and inferring treatment benefit at the subgroup level, a model can be trained to predict each patient’s recurrence risk with endocrine therapy alone and with chemoendocrine therapy. The difference between the two predictions is the patient’s estimated absolute benefit from chemotherapy.^13^

We applied this approach to develop Ataraxis Breast CTX, a multi-modal artificial intelligence (AI) test that estimates an individual patient’s absolute benefit from adjuvant chemotherapy directly from routine H&E-stained slides and clinical variables, without the need for additional genomic testing.^14^ Here, in a prespecified analysis approved by ECOG-ACRIN, we assessed the prognostic performance of CTX in the TAILORx trial, benchmarked against the prognostic performance of the RS. We also evaluated its predictive performance for chemotherapy benefit, testing whether it could identify chemosensitive patients missed by Oncotype DX.

## Methods

### Study design and population

TAILORx (NCT00310180) was a phase 3 randomized clinical trial that prospectively enrolled 10,273 women from April 2006 through October 2010, of whom 9719 were included in the primary intention-to-treat population. Eligible participants were women 18 to 75 years of age with HR+/HER2-, node-negative breast cancer, who were candidates for adjuvant chemotherapy according to NCCN guidelines, and consented to RS-based treatment assignment.^11^ Patients with low RS were assigned to endocrine therapy alone, and patients with high RS were assigned to chemoendocrine therapy. Patients with intermediate-RS were randomly assigned 1:1 to endocrine therapy alone or chemoendocrine therapy.

In our retrospective-prospective analysis of the TAILORx trial, we excluded 2984 of the 9719 patients (30.7%): 1716 had no slide, 477 did not pass quality control (low tissue area, incorrect staining, scanning artifacts, etc), 637 did not adhere to the assigned treatment (141 in the endocrine-therapy-alone group and 496 in the chemoendocrine group), 134 did not receive endocrine therapy, and 20 had missing outcome data (**Figure 1**). The final analytic cohort included 6735 patients, analyzed on a per-protocol basis. Slides were digitized by the ECOG-ACRIN Cancer Research Group with an Aperio AT2 scanner (Leica Biosystems) at ×20 magnification.

**Figure 1.**
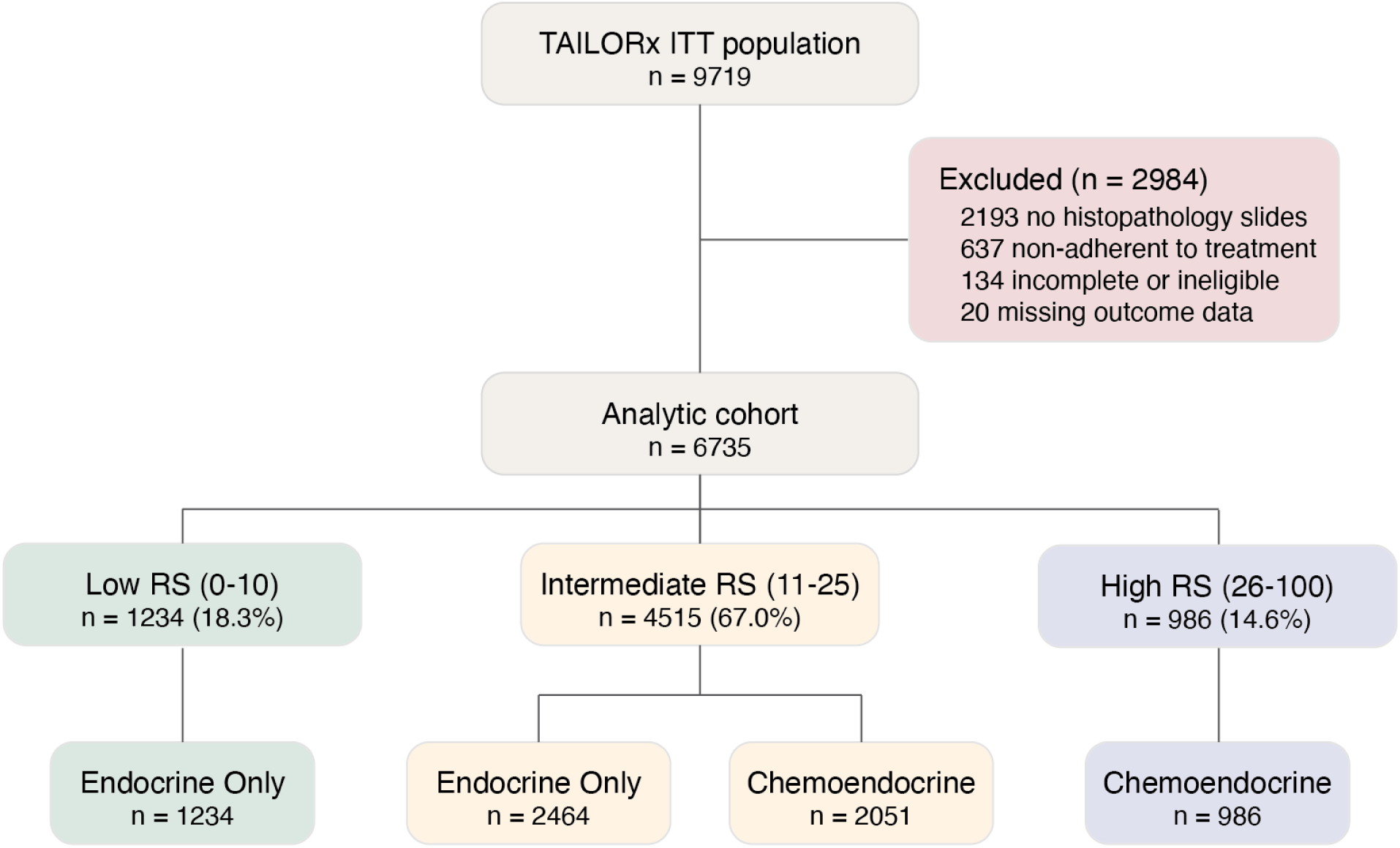
Patient Flow Diagram. Flowchart of patients from the TAILORx intention-to-treat population (N = 9719) through exclusions to the analytic cohort used for this analysis, stratified by treatment group. Patients in the analytic cohort were analyzed on a per-protocol basis. Ineligible patients include those who did not receive endocrine therapy. RS denotes Recurrence Score.

### Model development

Ataraxis Breast CTX is a causal multi-modal AI test that integrates morphologic features extracted from whole slide images of H&E-stained pathology slides with clinical variables to estimate individual chemotherapy benefit.^14^ CTX first analyzes digitized slides with a foundation model.^15^ CTX then estimates calibrated 5-year probabilities of recurrence under two treatment scenarios: endocrine therapy alone and chemoendocrine therapy. Estimated chemotherapy benefit is computed as the absolute reduction in predicted 5-year recurrence risk with chemoendocrine therapy, compared to endocrine therapy alone. CTX was trained on 12,821 patients from 21 independent cohorts. No data from TAILORx was used to train CTX. CTX was locked after development and not retrained or recalibrated for this study.

### Definition of counterfactual CTX scores

From the two recurrence predictions, two scores were derived. CTX-prognostic predicts the 5-year recurrence risk probability for each individual patient under the treatment they received. CTX-benefit is the difference between the two predicted probabilities, chemoendocrine therapy and endocrine therapy, and represents the estimated absolute reduction in recurrence probability attributable to chemotherapy.

### Study objective and end points

The primary objective of this analysis, prespecified in the protocol approved by ECOG-ACRIN, was to evaluate the prognostic performance of CTX-prognostic separately in patients who received endocrine therapy alone and in patients who received chemoendocrine therapy. Prognostic analyses included all four TAILORx treatment arms (arm A: low-RS assigned to endocrine therapy only, arm B: intermediate-RS randomized to endocrine therapy only, arm C: intermediate-RS randomized to chemoendocrine therapy, and arm D: high-RS assigned to chemoendocrine therapy). Prognostic analyses were stratified by treatment received. The secondary objective was to evaluate whether CTX-benefit is predictive of chemotherapy benefit, assessed by a treatment-by-CTX-benefit interaction test. This evaluation was performed in only patients with an intermediate-RS (arms B and C), where treatment was randomly assigned. Additional secondary analyses evaluated CTX-benefit in women aged 50 years or younger (in whom RS identified some chemotherapy benefit), and women over 50. In addition, CTX-prognostic’s prognostic performance was compared with the RS assay. The primary end point used was disease-free interval (DFI), defined as time to any recurrence event but not including death. Secondary end points of this analysis were recurrence-free interval (RFI), distant recurrence-free interval (DRFI), invasive disease-free survival (iDFS), and overall survival (OS). These secondary endpoints comprise the primary (iDFS) and secondary end points (RFI, DRFI, and OS) of the TAILORx trial.^11^ End point definitions are provided in **Table S1**. Analyses of secondary end points were not adjusted for multiplicity and are reported as hypothesis-generating.

### Statistical analysis

All analyses were performed in Python version 3.10.0. The lifelines,^16^ scikit-survival,^17^ scikit-learn,^18^ scipy,^19^ statsmodels,^20^ and patsy^21^ packages were used for survival modeling, performance metrics calculation, and statistical testing. Median follow up was estimated by the reverse Kaplan-Meier method.^22^ The prognostic performance of CTX-prognostic was assessed separately in patients treated with endocrine therapy alone and chemoendocrine therapy using C-index and time-dependent area under the receiver operating characteristic curve (AUROC) at 5 years. The proportional hazards assumption was tested using Schoenfeld residuals.^23^ As there was modest evidence of nonproportional hazards in the chemoendocrine group (p = 0.044), hazard ratios were interpreted as averages over the follow-up period. To assess whether CTX-prognostic contributes prognostic information beyond the RS, we fitted a multivariable Cox model including both scores as continuous predictors and reported the adjusted hazard ratio and p-value for CTX-prognostic. All Cox model hazard ratios are reported per 1 standard deviation (SD) increase in score. For univariate prognostic analyses of CTX-prognostic, hazard ratios were evaluated using one-sided Wald tests (α = 0.05, directional hypothesis: higher CTX-prognostic is associated with greater recurrence risk). One-sided tests were pre-specified as the alternative hypothesis was directional by design, where higher CTX-prognostic scores denote greater predicted recurrence risk. All other tests used a two-sided α of 0.05. Confidence intervals for C-index and time-dependent AUROC were estimated using 1000 bootstrap iterations.

The predictive performance of CTX-benefit was assessed in the intermediate-RS group. CTX-benefit was dichotomized at a threshold of 2%. This threshold was established and locked in prior validation studies of CTX.^14^ The predictive analysis fitted a Cox model that included treatment group, CTX-benefit, and an interaction term between them. Significance was tested by a likelihood ratio test against a nested model without the interaction. We constructed Kaplan-Meier curves for each treatment group within the high- and low-CTX-benefit categories and compared groups with a one-sided Wald test from an unadjusted Cox model. As prespecified, the interaction analysis was repeated in the subgroup of women aged 50 or under and over 50.

## Results

### Cohort details

The analytic cohort comprised 6735 patients with HR+/HER2-node-negative breast cancer enrolled in TAILORx (**Figure 1**). Baseline characteristics are described in **Table 1**. Of the 6735 patients, 1234 (18.3%) were classified as low-RS and received endocrine therapy alone; 4515 (67.0%) were classified as intermediate-RS, and were thus randomized for treatment receiving either endocrine therapy alone (n = 2464) or chemoendocrine therapy (n = 2051); 986 (14.6%) were classified as high-RS and received chemoendocrine therapy. The median follow-up was 7.3 years. A total of 353 DFI events were observed during follow-up.

**Table 1:** Baseline Characteristics of the Analytic Cohort by Treatment Group.

| Characteristic | Category | ET Only | CET |
| --- | --- | --- | --- |
| **N** |  | 3698 | 3037 |
| **Age** | < 50 | 962 | 867 |
|  | ≥ 50 | 2736 | 2170 |
| **T Stage** | Stage T1 | 2792 | 2162 |
|  | Stage T2 | 899 | 868 |
|  | Stage T3 | 6 | 5 |
|  | Stage T4 | 1 | 1 |
|  | Unknown | 0 | 1 |
| **Grade** | 1 | 1120 | 610 |
|  | 2 | 2077 | 1577 |
|  | 3 | 403 | 783 |
|  | Unknown | 98 | 67 |
| **CTX-Prognostic** | Median (IQR) | 0.059 (0.048-0.078) | 0.065 (0.054-0.084) |
| **CTX-Benefit** | CTX-Benefit-Low | 3378 | 2299 |
|  | CTX-Benefit-High | 320 | 738 |
| **21-RS Category** | Low- RS | 1234 | 0 |
|  | Intermediate- RS | 2464 | 2051 |
|  | High- RS | 0 | 986 |

CET denotes chemoendocrine therapy; ET, endocrine therapy alone.

### Recurrence risk stratification by CTX

CTX-prognostic was associated with DFI in both treatment groups. In univariate Cox models, CTX-prognostic showed hazard ratios of 1.651 (95% CI, 1.511-1.803, p < 0.001) and 1.642 (95% CI, 1.495-1.804, p < 0.001) in the endocrine and chemoendocrine groups, respectively. The C-index was 0.736 (95% CI, 0.697-0.770) in patients treated with endocrine therapy alone and 0.720 (95% CI, 0.681-0.756) in patients treated with chemoendocrine therapy (**Figure 2a, Table S3**). Time-dependent AUROC at 5 years was 0.765 (95% CI, 0.719-0.806) and 0.759 (95% CI, 0.712-0.800), respectively (**Figure 2a, Table S2**). Prognostic performance was similar across recurrence-based end points of RFI and DRFI and lower for iDFS and OS (**Figure S1**, **Table S3**).

**Figure 2.**
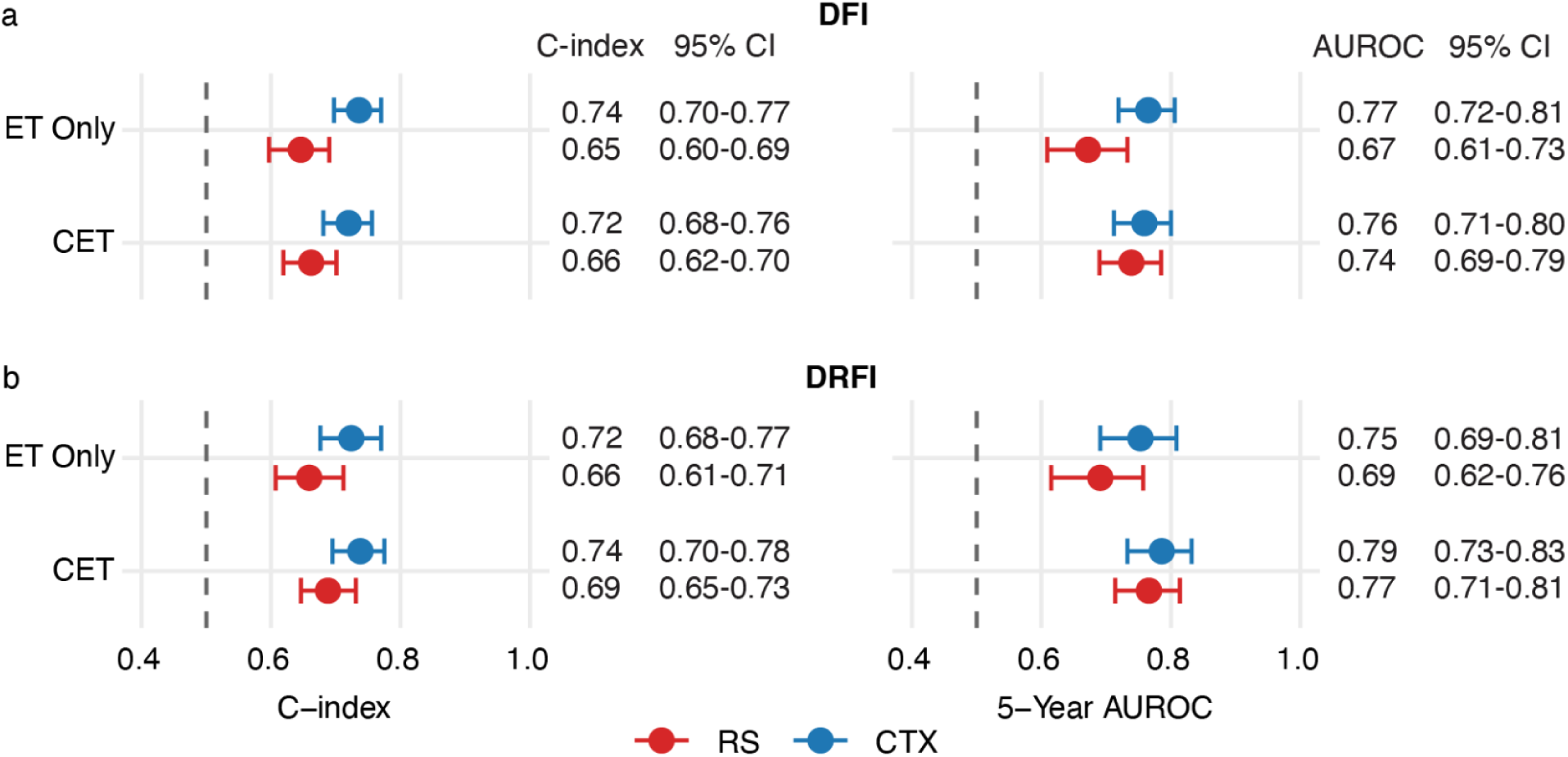
Prognostic Performance of CTX-Prognostic and the RS by Treatment Group Across All 6735 Patients. Panel a shows prognostic performance for the primary end point (DFI). The left plot shows the C-index for CTX-prognostic and the RS score in each treatment group. The right plot shows the 5-year time-dependent AUROC for CTX-prognostic and the RS score in each treatment group. Panel b shows the prognostic performance for the secondary end point DRFI where the left and right plots show C-index and 5-year AUROC respectively. Error bars represent 95% confidence intervals. CET denotes chemoendocrine therapy; CI, confidence interval; ET, endocrine therapy alone; and AUROC, area under the receiver operating curve.

### Identification of patients in the intermediate-RS population who benefit from chemotherapy

In the intermediate-RS population, 478 patients (10.6%) were classified as CTX-benefit-high and 4037 (89.4%) as CTX-benefit-low. Baseline characteristics of the intermediate-RS population, stratified by CTX-benefit categories, are summarized in **Table S3**. The effect of chemotherapy on disease-free interval significantly differed across CTX-benefit groups (p = 0.016; **Table 2)**, indicating that the effect of chemotherapy on DFI differed significantly between CTX-benefit categories. The treatment-by-benefit interaction for CTX was also significant across several secondary end points (RFI, DRFI, iDFS) but not for OS (**Table S4**).

**Table 2:**
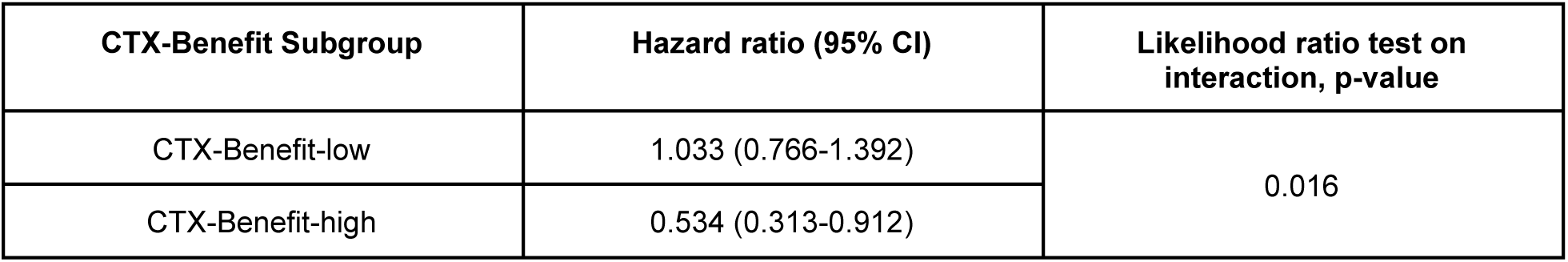
Cox Proportional Hazards Regression Model Assessing Interaction of CTX-Benefit Subgroup with Chemotherapy Benefit in RS-Intermediate Patients.

| CTX-Benefit Subgroup | Hazard ratio (95% CI) | Likelihood ratio test on interaction, p-value |
| --- | --- | --- |
| CTX-Benefit-low | 1.033 (0.766-1.392) | 0.016 |
| CTX-Benefit-high | 0.534 (0.313-0.912) |  |

In the primary TAILORx study, noninferiority of endocrine therapy alone compared with chemoendocrine therapy was reported.^11^ Consistent with this, in our analytical cohort, we did not detect a difference in DFI between treatment groups across the overall intermediate-RS population (HR = 0.89, 95% CI, 0.69-1.15, p = 0.368; **Figure S2a**). In the CTX-benefit-low category, DFI did not significantly differ between treatment groups (HR = 1.03, 95% CI, 0.77-1.39, p = 0.833, **Figure 3a**). The observed 5-year DFI in the endocrine therapy group was 97.8% (95% CI, 97.1-98.4%) and 97.8% (95% CI, 97.0-98.4%) in the chemoendocrine group, and 10-year DFI was 92.2% (95% CI, 89.4-94.3%) and 91.5% (95% CI, 87.9-94.0%), respectively. Thus, in the CTX low-benefit category, the absolute 5-year and 10-year benefit from the addition of chemotherapy were 0.0% and −0.7%, respectively, though 10-year estimates have high variance due to the small number of patients at risk. In contrast, in the CTX-benefit-high category, patients who were assigned to endocrine therapy only had significantly worse outcomes (HR = 0.53, 95% CI, 0.31-0.91, p = 0.011, **Figure 3b**). Their 5-year DFI was 89.6% (95% CI, 84.9-92.8%) compared with 95.2% for the patients in the chemoendocrine group (95% CI, 91.2-97.4%), corresponding to an absolute 5-year benefit of 5.6 percentage points from the addition of chemotherapy. At 10 years, DFI was 77.9% (95% CI, 69.3-84.4%) in patients treated with endocrine therapy only and 81.6% (95% CI, 66.2-90.4%) in patients treated with chemoendocrine therapy. Thus, the absolute 10-year benefit from the addition of chemotherapy was 3.7 percentage points.

**Figure 3.**
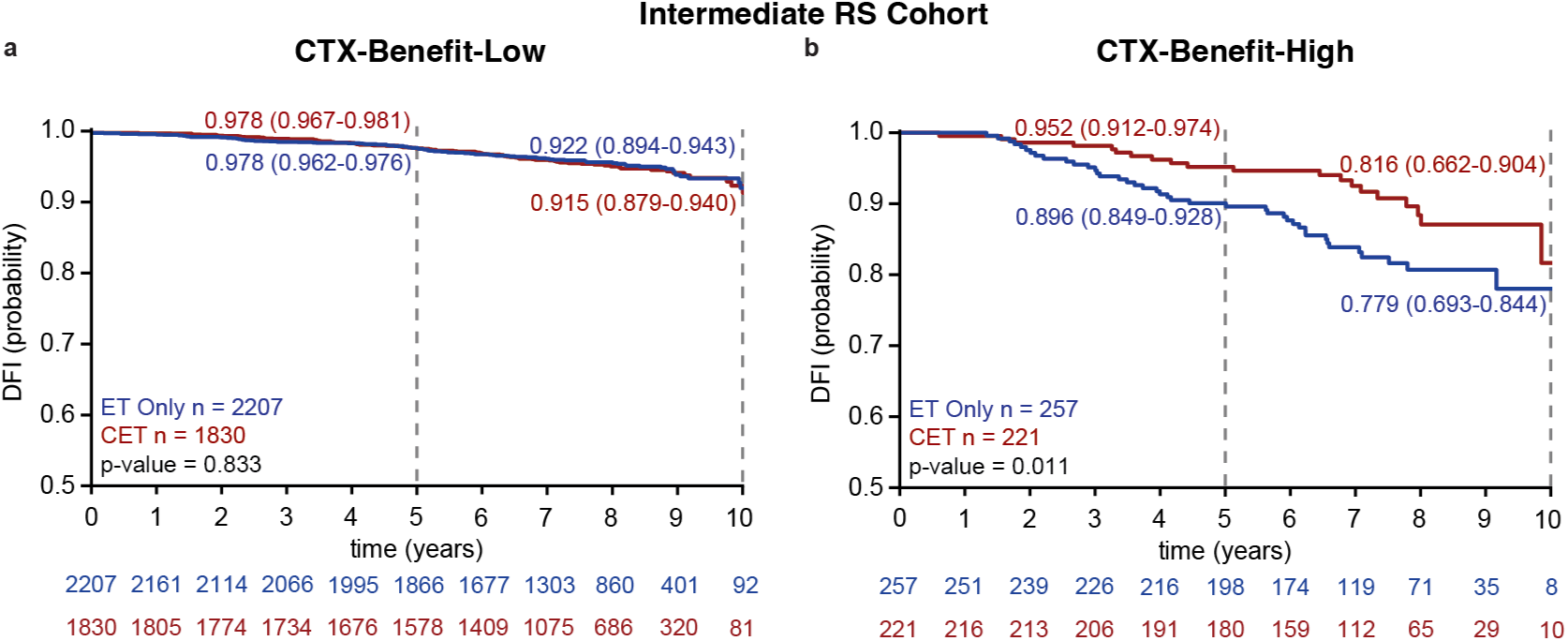
Kaplan-Meier Estimates of Disease-Free Interval by Treatment Group in the Intermediate-RS Population and CTX-benefit Subgroups. Panels a and b show DFI estimates in the overall intermediate-RS population that are CTX-benefit-low (panel a) and CTX-benefit-high (panel b). Five and ten-year DFI probabilities and 95% confidence intervals are shown for each group; p-values are derived from Wald tests of univariate Cox proportional hazards models. Numbers at risk at each time point are shown below each panel. CET denotes chemoendocrine therapy; CI, confidence interval; DFI, disease-free interval; ET, endocrine therapy alone; RS, Recurrence Score.

### CTX-benefit identifies chemotherapy benefit not captured by RS stratification

Correlating CTX-benefit with the RS, a moderate association was observed (Pearson’s r = 0.36), but CTX high-benefit patients were distributed across the RS range. Within the intermediate-RS population, the proportion classified as CTX-benefit-high rose from 3% at RS 11 to 26% at RS 25, and roughly two-thirds of all CTX-benefit-high patients had an RS of 20 or below (**Figure S3**).

To evaluate to what extent the treatment-effect heterogeneity identified by CTX-benefit is also captured by RS, we repeated the interaction analysis with the same cohort using RS rather than CTX-benefit. Using a threshold (RS ≥ 23) that yielded a high-RS group closest in size to the CTX-benefit high group (RS: n = 583, 13%; CTX-benefit: n = 478, 10.6%), the treatment-by-RS interaction was not statistically significant (p = 0.200, Figure S4, Table S5). In sensitivity analyses that treated both variables as continuous, the CTX-treatment-by-benefit interaction persisted (HR per 1 SD = 0.79, 95% CI, 0.70-0.90, p = 0.001) while the treatment-by-RS interaction was not significant (HR per 1 SD = 0.90, 95% CI, 0.69-1.19, p = 0.219).

### Age-stratified identification of women who benefit from chemotherapy

We evaluated the predictive performance of CTX-benefit among women with intermediate-RS, separately in those aged 50 years or younger and those older than 50 years. Among women 50 years of age or younger with high CTX-benefit, 5-year DFI was 86.0% with endocrine therapy alone and 96.6% with chemoendocrine therapy (p = 0.005; **Figure S5a**). DFI did not differ significantly between treatment groups in the CTX-benefit-low category (p = 0.077), whereas it was significantly longer with adjuvant chemotherapy in the CTX-benefit-high category (p = 0.005, **Figure S5b**). However, the treatment-by-CTX-benefit interaction was not significant in this subgroup (p = 0.083, **Table S6**). Among patients over 50, adjuvant chemotherapy was not associated with a significant difference in DFI in either CTX-benefit category (**Figure S5c**,**d**), and the treatment-by-CTX-benefit interaction was non-significant (p = 0.080, **Table S7**).

### CTX provides prognostic information beyond the RS

We assessed whether CTX added prognostic utility beyond the RS in the full cohort (n = 6735) by comparing performance for DFI and DRFI, the endpoints on which CTX and the RS, respectively, were developed and validated.^8,14^ CTX-prognostic yielded higher C-indexes than the RS for both end points in both treatment groups (**Figure 2** and **Table S3**). For DFI, the C-index was 0.74 (95% CI, 0.70-0.77) for CTX-prognostic and 0.65 (95% CI, 0.60-0.69) for the RS among patients who received endocrine therapy alone. In patients who received chemoendocrine therapy, the C-index for CTX-prognostic was 0.72 (95% CI, 0.68-0.76) compared to 0.66 (95% CI, 0.62-0.70). Similarly, for DRFI, CTX-prognostic had a C-index of 0.72 (95% CI, 0.68-0.77) and 0.74 (95% CI, 0.70-0.78) for endocrine therapy alone and chemoendocrine therapy respectively. In contrast, the C-index for RS was 0.66 (95% CI, 0.61-0.71) in patients treated with endocrine therapy alone and 0.69 (95% CI, 0.65-0.73) in patients treated with chemoendocrine therapy. Differences in C-index favored CTX-prognostic for DFI (p < 0.001 and p = 0.001 in the endocrine therapy alone and chemoendocrine groups, respectively) and for DRFI (p = 0.005 and p = 0.009).

For patients treated with endocrine therapy, 5-year AUROC was significantly higher for CTX-prognostic across both endpoints than RS for both DFI (CTX prognostic: 0.77, 95% CI, 0.72-0.81; RS: 0.67, 95% CI, 0.61-0.73; p < 0.001) and DRFI (CTX-prognostic: 0.75, 95% CI, 0.69-0.81; RS: 0.69, 95% CI, 0.62-0.76; p = 0.049). However, 5-year AUROC was not significantly different for patients treated with chemoendocrine therapy for DFI (CTX-prognostic: 0.76, 95% CI, 0.71-0.80; RS: 0.74, 95% CI, 0.69-0.79; p = 0.20) or DRFI (CTX-prognostic: 0.79, 95% CI, 0.73-0.83; RS: 0.77, 95% CI, 0.71-0.81; p = 0.19).

After adjustment for the RS, CTX-prognostic remained independently associated with DFI in both the endocrine therapy only group (HR = 1.576, 95% CI, 1.436-1.729, p < 0.001) and the chemoendocrine therapy group (HR = 1.540, 95% CI, 1.370-1.732, p < 0.001). The RS was also independently associated with DFI in both groups (endocrine therapy only: HR = 1.291, 95% CI, 1.096-1.521, p = 0.001; chemoendocrine therapy: HR = 1.151, 95% CI, 1.003-1.320, p = 0.023). Full results are listed in **Table S8**.

## Discussion

In the TAILORx trial, endocrine therapy alone was noninferior to chemoendocrine therapy for patients with an intermediate RS. However, no validated tool has been able to identify patients in this group whose tumors may be chemosensitive. In this analysis of the TAILORx, CTX, a causal multi-modal AI model, identified a subgroup comprising 11% of patients with an intermediate-RS in whom chemotherapy was associated with a 5% increase in 5-year DFI, whereas no benefit was detected among the remaining patients.

The RS was developed as a prognostic biomarker and has, at high scores, been associated with chemotherapy benefit^9,10,24^ but it does not estimate the benefit that an individual patient would receive from chemotherapy. In contrast, CTX provides two scores: CTX-prognostic, which estimates recurrence risk, and CTX-benefit, which estimates benefit from adjuvant chemotherapy. CTX-benefit-high patients were distributed across the intermediate-RS range (21.5% with RS 11-15, 44.4% with RS 16-20, 34.1% with RS 21-25), indicating that CTX-benefit identifies high-benefit patients who would not be captured by RS stratification alone.

To our knowledge, the present analysis is the first to validate in TAILORx, an individualized estimate of chemotherapy benefit developed entirely independently of the trial. Prior AI-based tools have relied on data from the trial: one paper trained directly on TAILORx data to predict RS scores rather than independent chemotherapy benefit,^25^ while another aimed to determine chemosensitivity but was trained and validated in low- and high-RS patients from TAILORx.^26^ Meanwhile, another multi-modal tool was developed independently of TAILORx and externally validated in the trial, but only for prognosis of late distant recurrence and not chemotherapy benefit.^27^

Independent validation in a retrospective analysis of a randomized clinical trial provides strong evidence for our test to be predictive, as treatment was randomly assigned and thus, treatment effects are isolated from confounding by indication.^28^ Our analysis adhered to the standard of a prospective-retrospective trial as the objectives were defined in a protocol approved by ECOG-ACRIN before any TAILORx outcome data were analyzed. In addition, the CTX model was locked before the analysis, no patients from TAILORx contributed to model training, and the CTX-benefit threshold was derived from independent development cohorts and applied without modification across validation studies.

Our study has limitations. First, complete histopathology slides were available for 77.4% of the TAILORx intention-to-treat population, and differential slide availability could introduce potential selection bias. However, the absolute standardized mean difference (averaged across clinical covariates) between the chemoendocrine arm and endocrine arm in the intermediate-RS subgroup was 0.06, suggesting that treatment confounding was negligible. Second, our analysis was restricted to node-negative disease and validation in randomized trials of node-positive disease is needed to determine whether CTX’s performance generalizes beyond the population studied here. Third, 637 patients were excluded because of non-adherence to assigned treatment, which may bias toward benefit. Fourth, the primary endpoint used in this study, DFI, does not align with a STEEP definition as it does not include breast cancer-specific mortality. This reflects a data limitation during model training, as cause of death was inconsistently recorded in real-world cohorts. Notably, the RFI endpoint used as a secondary endpoint does align with the STEEP definition, and does include breast-cancer-related deaths.

In conclusion, in the TAILORx trial, CTX was prognostic in patients treated with endocrine therapy alone and with chemoendocrine therapy, and its estimate of individual benefit was predictive of chemotherapy benefit among randomly assigned patients with an intermediate-RS. These findings suggest that individualized estimates of treatment benefit may refine chemotherapy decisions beyond population-averaged effects, but prospective evaluation is required before they are used in practice.

## Acknowledgements

Data was obtained from the TAILORx trial, conducted by ECOG-ACRIN (J.A. Sparano, V. Wang, R.J. Gray), with support from the NCI (U10CA180820 and U10CA180794).

## Data Availability

The data is not publicly available due to institutional and ethical constraints. Access to data from the TAILORx trial can be requested directly from ECOG-ACRIN.

## Code Availability

Ataraxis Breast CTX is available for non-commercial research use upon reasonable request. A Jupyter notebook to reproduce the analyses presented in this study is available upon request.

## Ethics Approvals

The TAILORx trial was conducted in accordance with the Declaration of Helsinki, the Belmont Report, and the U.S. Common Rule (45 CFR 46), and all participants provided written informed consent at enrollment. The present secondary analysis used retrospective, deidentified data and received approval from the WCG Institutional Review Board with a waiver of informed consent under 45 CFR 46.116(f). Data were obtained under an approved data access request and executed Data Use Certification from ECOG-ACRIN.

## Supplementary Appendix

This appendix has been provided by the authors to give readers additional information about their work.

Supplement to: Chan N, Tang C, Biswas D, et al. An AI Model Identifies Chemotherapy Benefit in Node-Negative HR+/HER2-Breast Cancer Patients from TAILORx, a Phase 3 Randomized Clinical Trial

## TAILORx Investigators

Joseph A. Sparano, M.D.^1^, Della F. Makower, M.D.^2^, Kathy S. Albain, M.D.^3^, Daniel F. Hayes, M.D.^4^, Charles E. Geyer, Jr., M.D.^5^, Elizabeth C. Dees, M.D.^5^, Matthew P. Goetz, M.D.^7^, John A. Olson, Jr., M.D.^8^, Sunil S. Badve, M.B.B.S., M.D.^9^, Thomas J. Saphner, M.D.^10^, Timothy J. Whelan, B.Sc., B.M., B.Ch., M.S.^11^, Virginia G. Kaklaman, M.D., D.Sc.^12^, Eleftherios P. Mamounas, M.D.^13^, Norman Wolmark, M.D.^14^

1. Icahn School of Medicine at Mount Sinai, Tisch Cancer Institute, New York, New York

2. Montefiore Medical Center, Albert Einstein College of Medicine, Bronx, New York

3. Loyola University Chicago Stritch School of Medicine, Maywood, Illinois

4. University of Michigan, Ann Arbor, Michigan

5. University of Pittsburgh, Pittsburgh, Pennsylvania

6. University of North Carolina, Chapel Hill, North Carolina

7. Mayo Clinic, Rochester, Minnesota

8. Washington University School of Medicine, St. Louis, Missouri

9. Winship Cancer Institute, Emory University, Atlanta, Georgia

10. Aurora Medical Center, Two Rivers, Wisconsin

11. McMaster University, Hamilton, Canada

12. University of Texas Health, San Antonio, Texas

13. AdventHealth Cancer Institute, Orlando, Florida

14. National Surgical Adjuvant Breast and Bowel Project, Pittsburgh, Pennsylvania

## Supplemental Figures

**Figure S1.**
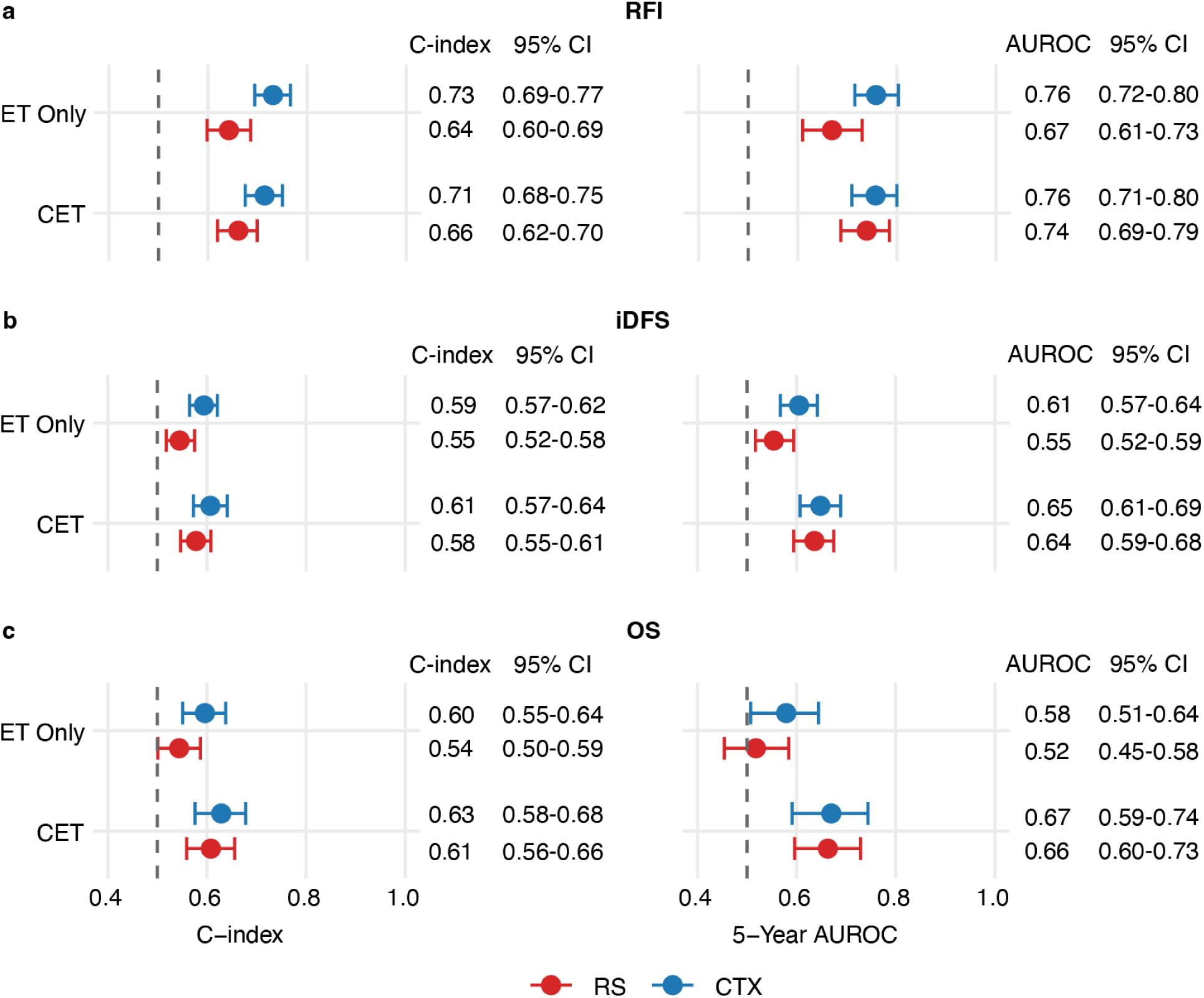
Prognostic Performance of CTX-Prognostic and the RS in Secondary End Points. Panel a shows prognostic performance for RFI. The left plot shows the C-index for CTX-prognostic and the RS score in each treatment group. The right plot shows the 5-year time-dependent AUROC. Panel b shows the prognostic performance for iDFS and panel c shows performance for OS. Error bars denote 95% confidence intervals. CET denotes chemoendocrine therapy; CI, confidence interval; and ET, endocrine therapy alone; AUROC, area under the receiver operating curve; RFI, recurrence-free interval; iDFS, invasive disease-free survival; and OS, overall survival.

**Figure S2.**
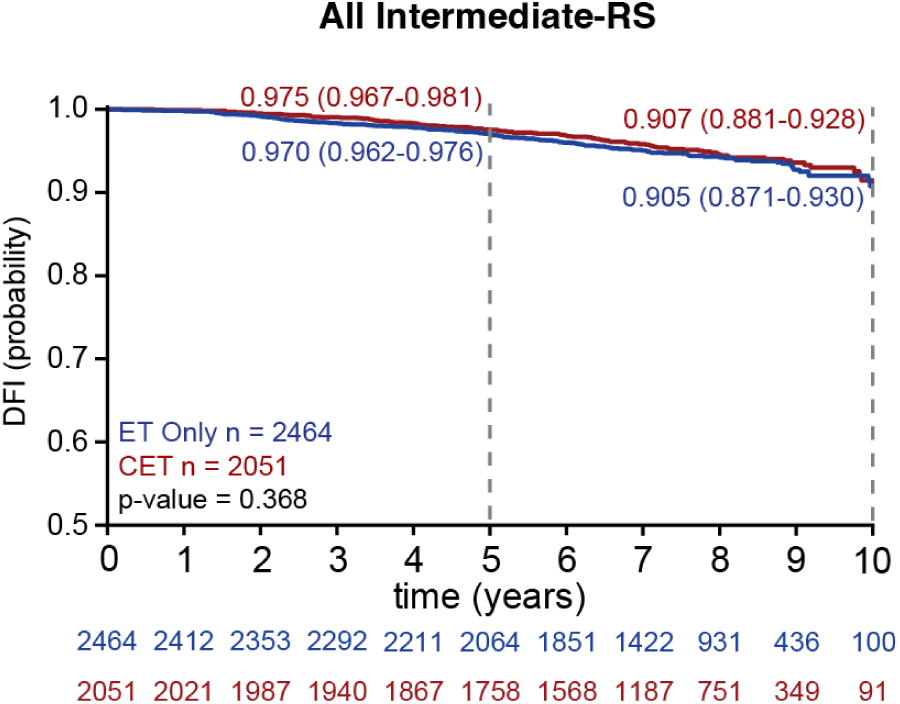
Kaplan-Meier Estimates of Disease-Free Interval by Treatment Group in the Overall Intermediate-RS Population. This figure shows DFI estimates in the overall intermediate-RS population. Five and ten-year DFI probabilities and 95% confidence intervals are shown for each group; p-values were derived from one-sided Wald tests of univariate Cox proportional hazards models. Numbers at risk at each time point are shown below each panel. CET denotes chemoendocrine therapy; CI, confidence interval; DFI, disease-free interval; ET, endocrine therapy alone.

**Figure S3.**
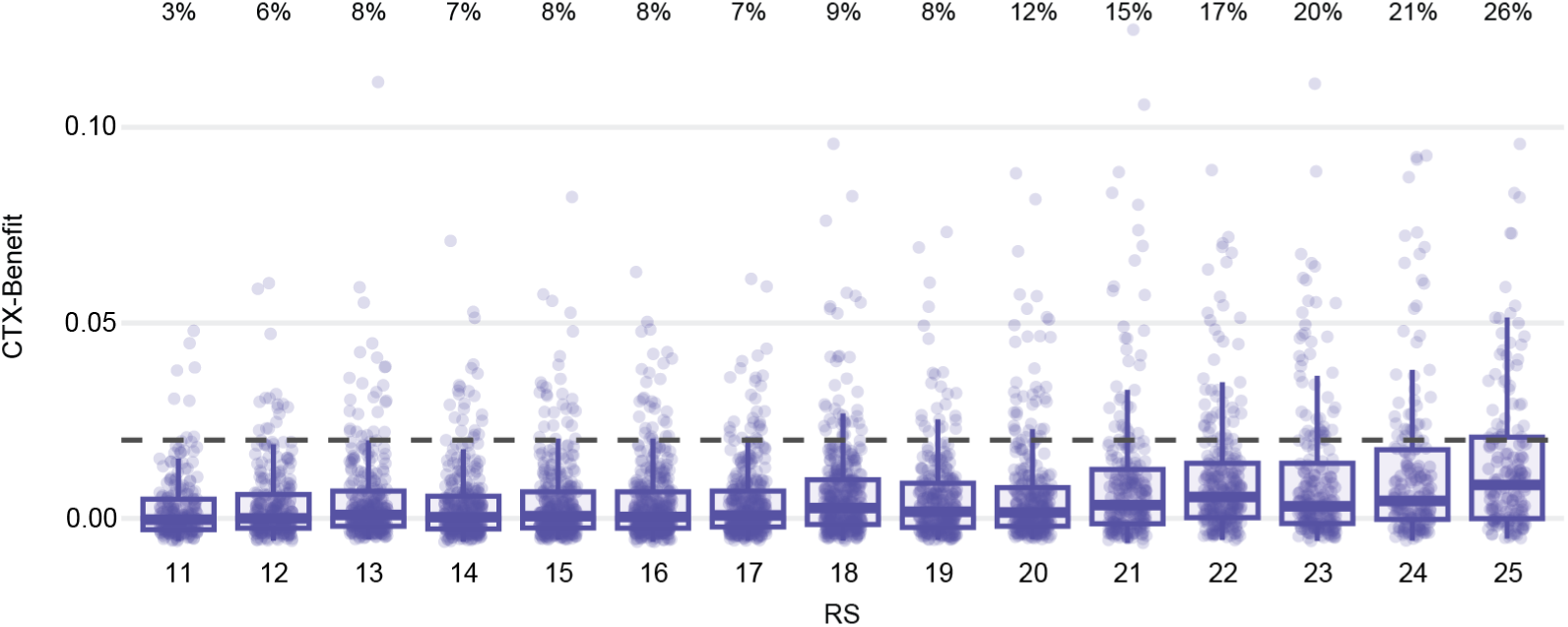
Joint Distribution of Recurrence Score and CTX-benefit. CTX-benefit values by individual RS in the intermediate-RS range (11-25); each point represents a single patient, the boxplots summarize the distribution of CTX-benefit at each RS, and the dashed horizontal line indicates the prespecified 2% threshold. Percentages above the boxplots indicate the proportion of patients at each RS classified as CTX-benefit-high. RS denotes Recurrence Score.

**Figure S4.**
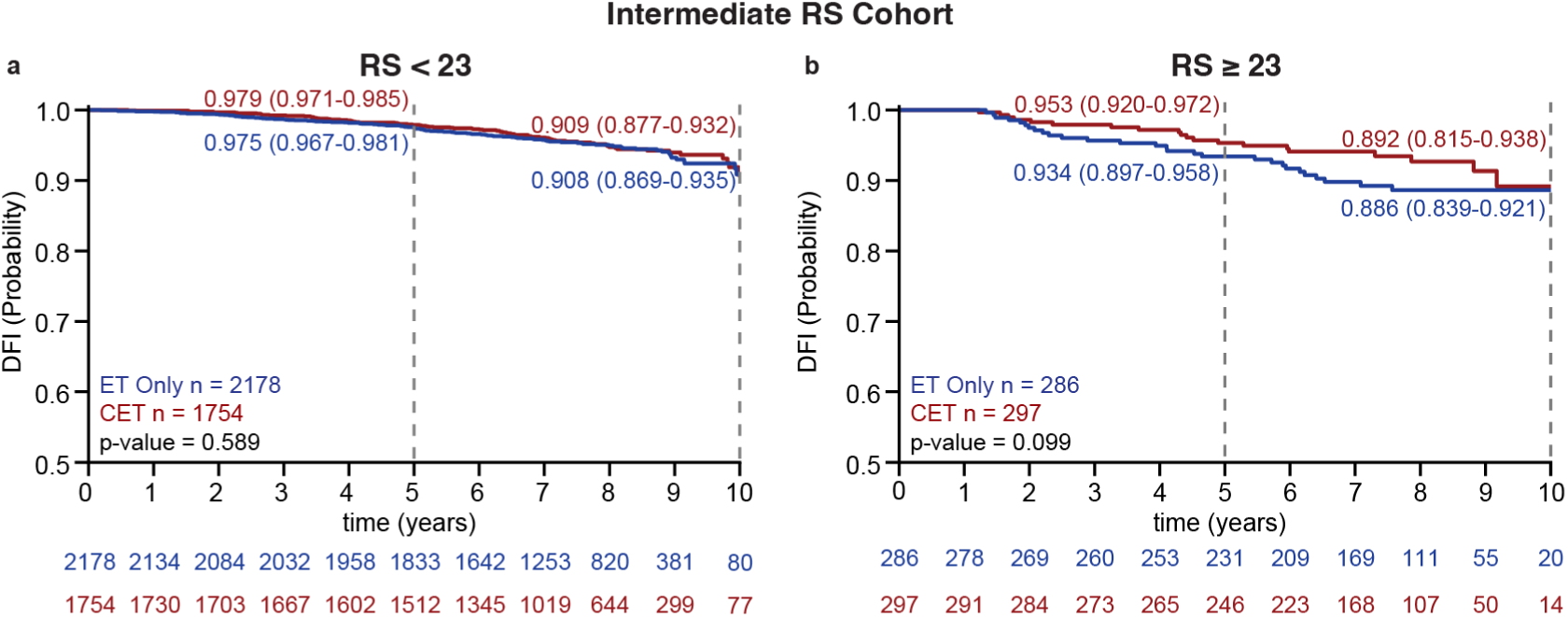
Kaplan-Meier Estimates of Disease-Free Interval by Treatment Group in the Intermediate-RS Population, Stratified by RS. Panel a shows DFI estimates in patients with an RS < 23 and panel b in patients with RS ≥ 23. Five- and ten-year DFI probabilities with 95% confidence intervals are shown for each group. Numbers at risk at each time point are shown below each panel. CET denotes chemoendocrine therapy; CI, confidence interval; DFI, disease-free interval; ET, endocrine therapy alone; and RS, Recurrence Score.

**Figure S5.**
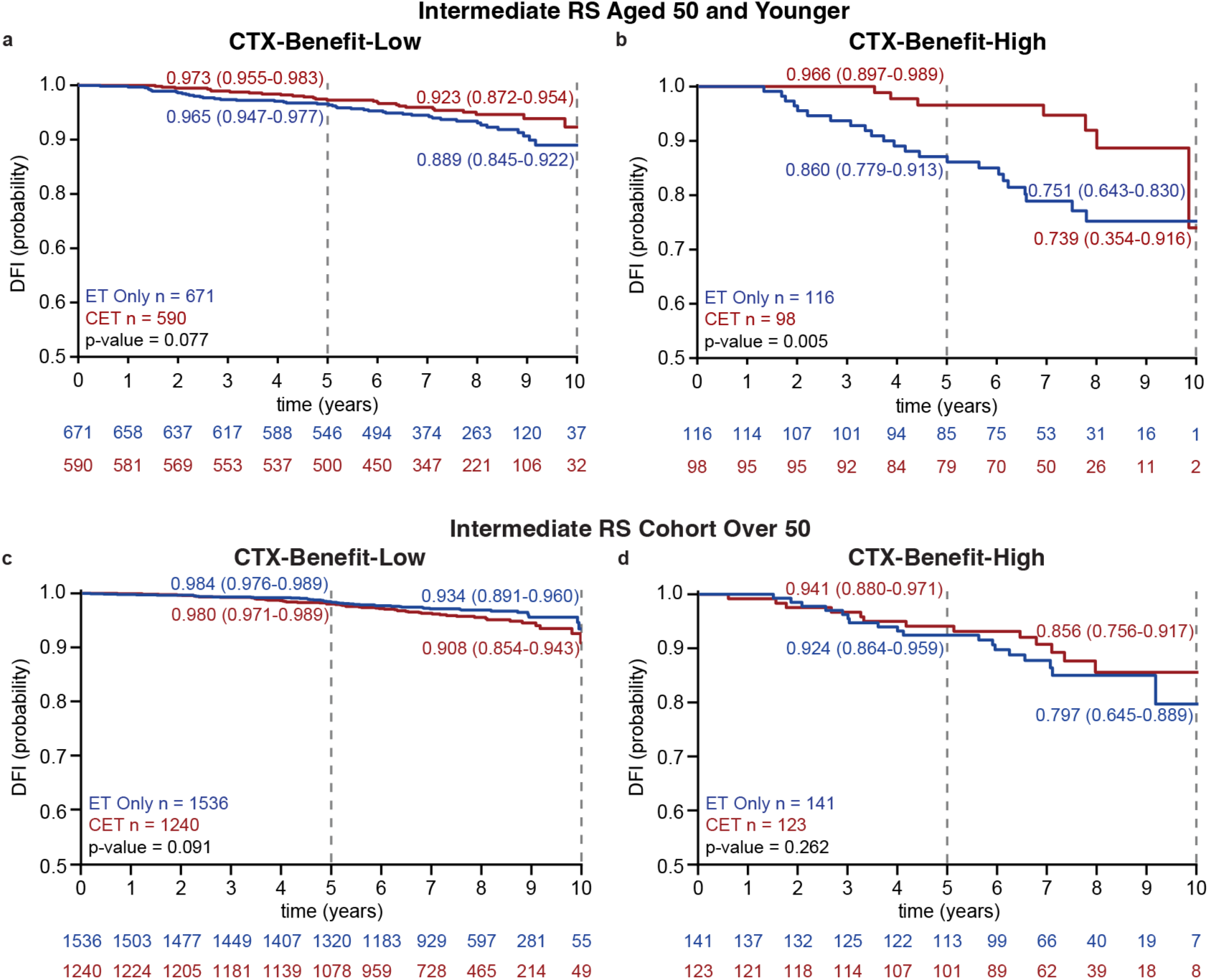
Kaplan-Meier Estimates of Disease-Free Interval by Treatment Group in the Intermediate-RS Population and CTX-Benefit Subgroups by Age. Panels a and b show the corresponding estimates in women aged 50 years or younger: CTX-benefit-low (panel a) and CTX-benefit-high (panel b), while panels c and d show estimates in women over 50. Five and ten-year DFI probabilities and 95% confidence intervals are shown for each group; p-values are calculated from Wald tests of univariate Cox proportional hazards models. Numbers at risk at each time point are shown below each panel. CET denotes chemoendocrine therapy; CI, confidence interval; DFI, disease-free interval; ET, endocrine therapy alone; RS, Recurrence Score.

## Supplemental Tables

**Table S1:** End Point Definitions.

|  | Invasive<br>IBTR | Local-Regional<br>Invasive<br>Recurrence | Distant<br>Recurrence | Death<br>from<br>Breast<br>Cancer | Death from<br>Non-Breast<br>Cancer | Death<br>from<br>Unknown<br>Cause | Invasive<br>Contralateral<br>Breast<br>Cancer | Second<br>Primary<br>Invasive<br>Cancer |
| --- | --- | --- | --- | --- | --- | --- | --- | --- |
| **DFI** | ✓ | ✓ | ✓ |  |  |  |  |  |
| **RFI** | ✓ | ✓ | ✓ | ✓ |  |  |  |  |
| **DRFI** |  |  | ✓ | ✓ |  |  |  |  |
| **iDFS** | ✓ | ✓ | ✓ | ✓ | ✓ | ✓ | ✓ | ✓ |
| **OS** |  |  |  | ✓ | ✓ | ✓ |  |  |

DFI denotes disease-free interval; RFI, recurrence-free interval; DRFI, distant recurrence-free interval; iDFS, invasive disease-free survival; and OS, overall survival.

**Table S2:** Prognostic Performance of CTX-Prognostic and the RS Across Primary and Secondary End Points by Treatment Group.

| End Point | Group | Metric | CTX-Prognostic | RS |
| --- | --- | --- | --- | --- |
| DFI | ET | C-index | 0.736 (0.697-0.770) | 0.646 (0.597-0.690) |
|  |  | 5-year AUROC | 0.765 (0.719-0.806) | 0.672 (0.609-0.733) |
|  | CET | C-index | 0.720 (0.681-0.756) | 0.662 (0.619-0.701) |
|  |  | 5-year AUROC | 0.759 (0.712-0.800) | 0.739 (0.690-0.785) |
| RFI | ET | C-index | 0.731 (0.694-0.766) | 0.642 (0.598-0.680) |
|  |  | 5-year AUROC | 0.758 (0.715-0.803) | 0.669 (0.610-0.730) |
|  | CET | C-index | 0.714 (0.675-0.750) | 0.661 (0.619-0.699) |
|  |  | 5-year AUROC | 0.757 (0.709-0.800) | 0.739 (0.687-0.785) |
| iDFS | ET | C-index | 0.594 (0.565-0.621) | 0.545 (0.518-0.575) |
|  |  | 5-year AUROC | 0.605 (0.567-0.642) | 0.554 (0.517-0.594) |
|  | CET | C-index | 0.607 (0.573-0.641) | 0.578 (0.547-0.608) |
|  |  | 5-year AUROC | 0.648 (0.607-0.689) | 0.636 (0.594-0.675) |
| DRFI | ET | C-index | 0.724 (0.676-0.770) | 0.659 (0.607-0.712) |
|  |  | 5-year AUROC | 0.753 (0.691-0.809) | 0.692 (0.615-0.757) |
|  | CET | C-index | 0.738 (0.695-0.775) | 0.688 (0.646-0.731) |
|  |  | 5-year AUROC | 0.786 (0.733-0.832) | 0.766 (0.714-0.814) |
| OS | ET | C-index | 0.596 (0.551-0.638) | 0.544 (0.501-0.587) |
|  |  | 5-year AUROC | 0.579 (0.507-0.644) | 0.518 (0.454-0.584) |
|  | CET | C-index | 0.629 (0.576-0.678) | 0.608 (0.559-0.656) |
|  |  | 5-year AUROC | 0.670 (0.591-0.744) | 0.663 (0.596-0.729) |

DFI denotes disease-free interval; RFI, recurrence-free interval; DRFI, distant recurrence-free interval; iDFS, invasive disease-free survival; OS, overall survival; ET, endocrine therapy alone; CET, chemoendocrine therapy; and AUROC, area under the receiver operating characteristic curve.

**Table S3:** Patient Characteristics in RS-Intermediate Patients by CTX-Benefit Group.

| Characteristic | Category | CTX-Benefit-Low | CTX-Benefit-High |
| --- | --- | --- | --- |
| **N** |  | 4037 | 478 |
| **Age (p < 0.001)** | ≤ 50 | 1261 (27.1%) | 214 (41.4%) |
|  | > 50 | 2776 (72.9%) | 264 (58.6%) |
| **T Stage (p < 0.001)** | Stage T1 | 3261 (80.8%) | 124 (26.0%) |
|  | Stage T2 | 772 (19.1%) | 349 (73.0%) |
|  | Stage T3 | 4 (0.1%) | 4 (0.8%) |
|  | Stage T4 | 0 (0%) | 1 (0.2%) |
| **Grade (p < 0.001)** | 1 | 1237 (30.6%) | 43 (9.0%) |
|  | 2 | 2241 (55.5%) | 264 (55.2%) |
|  | 3 | 461 (11.4%) | 156 (32.7%) |
|  | Unknown | 98 (2.5%) | 15 (3.1%) |
| **Treatment received (p = 0.73)** | Endocrine only | 2207 (54.7%) | 257 (53.8%) |
|  | Chemoendocrine | 1830 (45.3%) | 221 (46.2%) |
| **CTX Prognostic (p < 0.001)** | Median (IQR) | 0.057 (0.049-0.070) | 0.116 (0.097-0.142) |

NOTE. Data are presented as No. (%) unless otherwise indicated; percentages are column percentages calculated within each CTX-benefit group. Fisher exact test was used to calculate p-values for categorical variables (age category, T stage, grade, and treatment received) were compared to determine if distributions differed between CTX-benefit groups with Fisher's exact tests; the test for grade was performed on patients with known grade only, excluding those with unknown grade. The p-value for the CTX-prognostic score, expressed as median (IQR), was calculated using the Wilcoxon rank-sum test.

**Table S4:** Cox Proportional Hazards Models Assessing Interaction of CTX-Benefit Subgroup with Chemotherapy Benefit in RS-Intermediate Patients Across Secondary End Points.

| End Point | CTX-Benefit category | Hazard ratio (95% CI) | p-value |
| --- | --- | --- | --- |
| RFI | Low | 1.053 (0.788-1.406) | 0.013 |
|  | High | 0.534 (0.313-0.912) |  |
| iDFS | Low | 0.895 (0.748-1.070) | 0.015 |
|  | High | 0.513 (0.327-0.804) |  |
| DRFI | Low | 1.243 (0.869-1.778) | 0.004 |
|  | High | 0.483 (0.258-0.905) |  |
| OS | Low | 0.990 (0.746-1.312) | 0.054 |
|  | High | 0.531 (0.267-1.058) |  |

RFI denotes recurrence-free interval; DRFI, distant recurrence-free interval; iDFS, invasive disease-free survival; and OS, overall survival.

**Table S5:** Cox Proportional Hazards Regression Model in RS-Intermediate Patients, Assessing Interaction of RS Subgroup with Chemotherapy Benefit.

| RS Subgroup | Hazard ratio (95% CI) | Likelihood ratio test on interaction, p-value |
| --- | --- | --- |
| RS < 23 | 0.923 (0.691-1.234) | 0.200 |
| RS ≥ 23 | 0.686 (0.386-1.217) |  |

**Table S6:** Cox Proportional Hazards Models Assessing Interaction of CTX-Benefit Subgroup with Chemotherapy Benefit in RS-Intermediate Patients Aged 50 or Younger.

| End Point | CTX-Benefit subgroup | Hazard ratio (95% CI) | p-value |
| --- | --- | --- | --- |
| DFI | Low | 0.651 (0.405-1.047) | 0.083 |
|  | High | 0.331 (0.142-0.771) |  |

DFI denotes disease-free interval; RFI, recurrence-free interval; DRFI, distant recurrence-free interval; iDFS, invasive disease-free survival; OS, overall survival; and Inf, infinity.

**Table S7:**
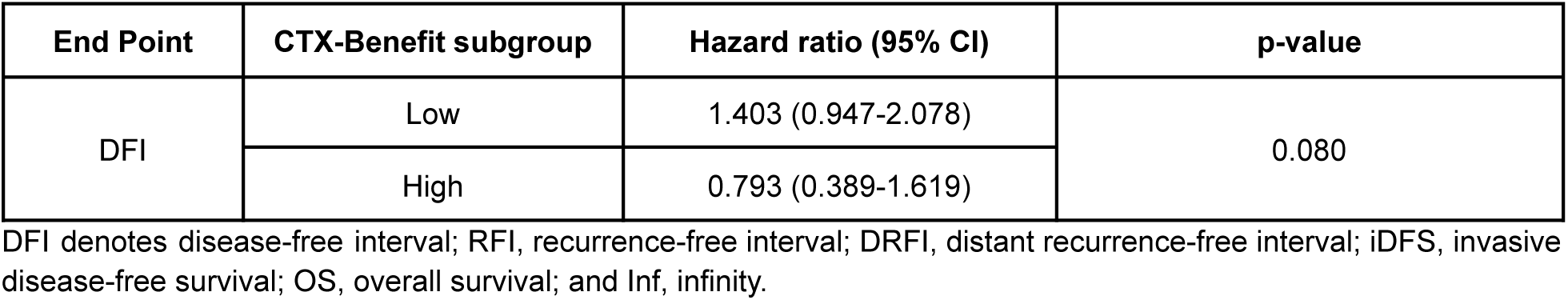
Cox Proportional Hazards Models Assessing Interaction of CTX-Benefit Subgroup with Chemotherapy Benefit in RS-Intermediate Patients Over 50.

| End Point | CTX-Benefit subgroup | Hazard ratio (95% CI) | p-value |
| --- | --- | --- | --- |
| DFI | Low | 1.403 (0.947-2.078) | 0.080 |
|  | High | 0.793 (0.389-1.619) |  |

DFI denotes disease-free interval; RFI, recurrence-free interval; DRFI, distant recurrence-free interval; iDFS, invasive disease-free survival; OS, overall survival; and Inf, infinity.

**Table S8:** Multivariable Cox Proportional Hazards Models Including CTX-Prognostic and the RS.

| Group | Score | DFI |  | DRFI |  |
| --- | --- | --- | --- | --- | --- |
|  |  | Hazard Ratio | p-value | Hazard Ratio | p-value |
| ET | CTX-Prognostic | 1.576 (1.436-1.729) | < 0.001 | 1.603 (1.437-1.788) | < 0.001 |
|  | RS | 1.291 (1.096-1.521) | 0.001 | 1.330 (1.091-1.621) | 0.002 |
| CET | CTX-Prognostic | 1.540 (1.370-1.732) | < 0.001 | 1.578 (1.390-1.790) | < 0.001 |
|  | RS | 1.151 (1.003-1.320) | 0.023 | 1.206 (1.042-1.395) | 0.006 |

DFI denotes disease-free interval; DRFI, distant recurrence-free interval; ET, endocrine therapy alone; and CET, chemoendocrine therapy.

## References

1. Howlader, N. et al. US incidence of breast cancer subtypes defined by joint hormone receptor and HER2 status. J. Natl. Cancer Inst. 106, dju055 (2014).

2. Fisher, B. et al. Tamoxifen and chemotherapy for lymph node-negative, estrogen receptor-positive breast cancer. J. Natl. Cancer Inst. 89, 1673–1682 (1997).

3. Albain, K. S. et al. Adjuvant chemotherapy and timing of tamoxifen in postmenopausal patients with endocrine-responsive, node-positive breast cancer: a phase 3, open-label, randomised controlled trial. Lancet 374, 2055–2063 (2009).

4. Early Breast Cancer Trialists’ Collaborative Group (EBCTCG). Aromatase inhibitors versus tamoxifen in early breast cancer: patient-level meta-analysis of the randomised trials. Lancet 386, 1341–1352 (2015).

5. Smith, G. L. et al. Financial burdens of cancer treatment: A systematic review of risk factors and outcomes. J. Natl. Compr. Canc. Netw. 17, 1184–1192 (2019).

6. Zafar, S. Y. & Abernethy, A. P. Financial toxicity, Part I: a new name for a growing problem. Oncology (Williston Park) 27, 80–1, 149 (2013).

7. Shapiro, C. L. & Recht, A. Side effects of adjuvant treatment of breast cancer. N. Engl. J. Med. 344, 1997–2008 (2001).

8. Paik, S. et al. A multigene assay to predict recurrence of tamoxifen-treated, node-negative breast cancer. N. Engl. J. Med. 351, 2817–2826 (2004).

9. Albain, K. S. et al. Prognostic and predictive value of the 21-gene recurrence score assay in postmenopausal women with node-positive, oestrogen-receptor-positive breast cancer on chemotherapy: a retrospective analysis of a randomised trial. Lancet Oncol. 11, 55–65 (2010).

10. Geyer, C. E., Jr et al. 21-Gene assay as predictor of chemotherapy benefit in HER2-negative breast cancer. NPJ Breast Cancer 4, 37 (2018).

11. Sparano, J. A. et al. Adjuvant chemotherapy guided by a 21-gene expression assay in breast cancer. N. Engl. J. Med. 379, 111–121 (2018).

12. Kwa, M., Makris, A. & Esteva, F. J. Clinical utility of gene-expression signatures in early stage breast cancer. Nat. Rev. Clin. Oncol. 14, 595–610 (2017).

13. Künzel, S. R., Sekhon, J. S., Bickel, P. J. & Yu, B. Metalearners for estimating heterogeneous treatment effects using machine learning. Proc. Natl. Acad. Sci. U. S. A. 116, 4156–4165 (2019).

14. Biswas, D., et al. Causal multi-modal AI for personalized chemosensitivity prediction. *arXiv* (2026).

15. Cappadona, J. et al. Squeezing performance from pathology foundation models with chained hyperparameter searches. in NeurIPS 2024 Workshop: Self-Supervised Learning - Theory and Practice (2024).

16. Davidson-Pilon, C. lifelines: survival analysis in Python. Journal of Open Source Software 4, 1317 (2019).

17. Pölsterl, S. scikit-survival: A Library for Time-to-Event Analysis Built on Top of scikit-learn. Journal of Machine Learning Research 21, 1–6 (2020).

18. Pedregosa, F. et al. Scikit-learn: Machine Learning in Python. Journal of Machine Learning Research 12, 2825–2830 (2011).

19. Virtanen, P. et al. SciPy 1.0: fundamental algorithms for scientific computing in Python. Nat. Methods 17, 261–272 (2020).

20. Seabold, S. & Perktold, J. Statsmodels: Econometric and statistical modeling with python. in Proceedings of the Python in Science Conference 92–96 (SciPy, 2010).

21. Patsy: Describing Statistical Models in Python Using Symbolic Formulas. (Github).

22. Schemper, M. & Smith, T. L. A note on quantifying follow-up in studies of failure time. Control. Clin. Trials 17, 343–346 (1996).

23. Grambsch, P. M. & Therneau, T. M. Proportional hazards tests and diagnostics based on weighted residuals. Biometrika 81, 515–526 (1994).

24. Paik, S. et al. Gene expression and benefit of chemotherapy in women with node-negative, estrogen receptor-positive breast cancer. J. Clin. Oncol. 24, 3726–3734 (2006).

25. Shamai, G. et al. Deep learning on histopathological images to predict breast cancer recurrence risk and chemotherapy benefit: a multicentre, model development and validation study. Lancet Oncol. 0, (2026).

26. Piven, A. et al. Predicting chemotherapy benefit in premenopausal women with intermediate genomic scores using deep learning. J. Clin. Oncol. 44, 602–602 (2026).

27. Mamounas, E. P. et al. Development and validation of a multimodal-multitask deep learning approach for estimating late distant recurrence risk in HR-positive early breast cancer. Cancer Res. Commun. 6, 1825–1835 (2026).

28. Simon, R. M., Paik, S. & Hayes, D. F. Use of archived specimens in evaluation of prognostic and predictive biomarkers. J. Natl. Cancer Inst. 101, 1446–1452 (2009).

